# Videographic Analysis of an Intervention to Improve Patient-Centered Care for People Living with Type 2 Diabetes: the QBSAFE Randomized Trial

**DOI:** 10.64898/2026.02.06.26345767

**Authors:** Victor V Montori, Felipe Larios, Satya Sai Sri Bandi, Ana Cristina Proano, Kerly Guevara, Luis Vilatuna, Shubhangi Bagewadi, Anka van Gastel, Megan Branda, Anne Camp, Mel Montosa, Rozalina McCoy, Victor M Montori, Kasia J Lipska

**Affiliations:** Knowledge and Evaluation Research Unit, Mayo Clinic, Rochester, MN, U.S.A.; LSU Health Shreveport, Shreveport, LA, U.S.A.; Leiden University Medical Center, Leiden, Netherlands; Fair Haven Community Health Center, New Haven, CT, U.S.A.; University of Maryland School of Medicine, Baltimore, MD, U.S.A.; University of Maryland Institute for Health Computing, North Bethesda, MD, U.S.A.; Yale School of Medicine, New Haven, CT, U.S.A.

## Abstract

**Background:** The self-management of type 2 diabetes (T2D) typically requires enacting various lifestyle changes, which can challenge people living with T2D. Clinical encounters between people with T2D and their clinicians, however, are often focused on metabolic management, leaving less time available for other self-management topics. The QBSAFE cards help patients articulate aspects of their experience with diabetes and prioritize issues for discussion.

**Methods:** This report details secondary outcomes of a randomized controlled trial; primary outcomes are reported elsewhere. All data was collected at Fair Haven Community Health Care, a federally qualified primary care clinic. 11 clinicians were randomly assigned to provide either usual care or usual care with QBSAFE cards to 155 of their patients with type 2 diabetes and hemoglobin A1c >8%. All patient encounters were video recorded for analysis. Patients and clinicians were not blinded to arm allocation but were kept unaware of the specific aims of the trial. Encounter video reviewers were blinded to arm allocation, but not to specific aims of the trial. The outcomes of interest for this report were the extent to which the QBSAFE cards were used as intended, their effect on the topics of discussion, and whether they enabled clinicians to notice and respond to each patient’s situation; comparisons between arms were conducted by a linear mixed model with fixed effect of arm and cluster effect of clinician, analyzed in both intent-to-treat and per-protocol populations.

**Findings:** 12 patients were excluded post-randomization (A1c <8%). Of 143 eligible patients, 137 encounters (65 in the usual care arm, 72 in QBSAFE) yielded evaluable videos. QBSAFE was used as intended in 61 (85%) QBSAFE arm encounters. Conversations about burden of treatment related to non-pharmacological interventions (17 vs 33, *p*= 0·04) and taking medications (11 vs 33, *p*= 0·0008) and about the patient’s challenging environment (2 vs 10, *p*= 0·04) were more prevalent in the QBSAFE group. There was no difference in the rate of conversations about metabolic management or of new care plans as a result of conversations between groups.

**Interpretation:** While there was a difference in the types of conversations observed between the two study arms, this difference was small and only apparent in a few domains. Future work could aim to modify the QBSAFE cards to more effectively stimulate patient-centered discussions and to further prepare clinicians to respond to a variety of issues raised during the clinical visit.

**Funding:** This work was supported by funding from the National Institute of Diabetes and Digestive and Kidney Diseases (R01DK129616).

## Introduction

The self-management of type 2 diabetes (T2D) typically requires enacting lifestyle changes, performing glycemic self-monitoring, the ongoing use of one or more glucose lowering agents, and attending regular clinical follow-ups with laboratory tests and clinical visits. These demands can challenge people living with T2D and affect how well they can live their lives. Their situation is almost always complicated by other health conditions, each with its own self-care demands^1^ and other pressures related to family, work, and community responsibilities. Clinical encounters between people with T2D and their clinicians, however, are often dominated by focus on metabolic management (changes in hemoglobin A1C [HbA1c] or other indicators of glycemia) leaving less time available for discussion of other self-management topics that may be more important to people with T2D. It is desirable for clinicians to identify, discuss, and address the full range of patients’ care needs and priorities, enabling the co-creation of care plans that align with the patient’s preferences, life routines, and clinical and personal needs.^2^

Interventions within the clinical encounter have previously succeeded in facilitating discussions between patients and clinicians.^3^ For example, shared decision-making tools that display treatment options and their features can help clinicians and patients engage in meaningful discussions about choice of glucose-lowering pharmacotherapy.^4^ Their deployment assumes that the patient’s concerns to be addressed and the options to address them are already known; a more patient-centered approach would require taking a step back to first determine what, specifically, should be discussed.

The QBSAFE cards are a series of cards that reflect aspects of the QBSAFE domains (Quality of life, Burden of treatment, Safety, Avoidance of Future Events),^5^ that help patients articulate aspects of their experience with diabetes that are important to them and that they would like to prioritize for discussion. The patient selects up to three cards and brings them into the consultation for review with their clinician. A randomized trial of usual care with QBSAFE cards vs. usual care alone has reported their impact on clinical outcomes elsewhere.

This report addresses the extent to which the QBSAFE cards were used as intended, also called fidelity of use, in the intervention arm; whether they were used in the control arm (i.e., contamination), what effect their use had on the topics of discussion, and whether their use enabled clinicians to notice and respond to each patient’s situation.

## METHODS

### Study design

We performed a videographic analysis of 143 recorded patient-clinician encounters collected for a cluster randomized trial to assess the efficacy of the QBSAFE cards (NCT05553912). The primary results of this trial, focused on efficacy with respect to glycemic management and illness intrusiveness, are reported elsewhere. This is a secondary, pre-specified analysis. The CONSORT checklist is available in Supplemental Material 1. All study procedures were approved by the Mayo Clinic IRB (22-007315) and Yale IRB (#2000032654).

### Setting

All data was collected at Fair Haven Community Health Care, a federally qualified primary care clinic. This medical center cares for a diverse population of English and Spanish speakers; of the over 30,000 patients receiving care at Fair Haven annually, over 1,500 adults live with T2D.

### Intervention

There are 14 QBSAFE cards addressing issues related to quality of life, burden of treatment, safety, and prevention of diabetes complications. The set is available in both English and Spanish and in three different formats: as a single printed sheet, as 14 discrete cards, or as an interactive website (https://patientrevolution.org/qbsafe).^5^

### Participants

Eligible clinicians included any adult primary care physician, nurse practitioner, or physician assistant participating in the care of and prescribing medications for patients with T2D. Eligible patients included adults (age ≥18 years) diagnosed with T2D, able to give written informed consent to participate in the trial, fluent in either English or Spanish, and with an HbA1c >8% at the study start date. Patients for whom an HbA1c target >8% is clinically reasonable (e.g., those with limited life expectancy) and/or aligned with goals/preferences were excluded. No changes to eligibility criteria were made after trial started.

### Randomization and blinding

Randomization was at the clinician level to minimize contamination. Participating clinicians were randomly allocated on a 1:1 ratio via computer-generated sequence using the Pocock-Simon method to either usual care (control arm) or usual care with the QBSAFE cards (QBSAFE arm) minimizing imbalance across clinician site and fluency in Spanish. Clinicians learned of their allocation after they had given consent to participate in the trial. Clinicians in the QBSAFE arm attended a training session where researchers presented the cards with their intended use and were provided materials presenting suggested ways of responding to some of the cards. Although blinding participants to trial arm was not feasible, patients and clinicians were kept unaware of the specific aims of the trial. Video encounter reviewers were blinded to arm allocation, but not to specific aims of the trial.

### Recruitment/Data Collection

A trained study coordinator recruited patients over the phone, by mail, email, or by patient portal message. Patients were asked to arrive early to their scheduled appointment, where the consent process was completed in a private exam room before their appointment. Patients assigned to the intervention group were introduced to the QBSAFE cards in the waiting area or in the exam room and were asked to choose up to 3 cards to discuss during the clinical encounter. Clinical encounters were video recorded. Participants could turn the camera off at any point or direct its lens to the ceiling to capture only audio. Incomplete encounter recordings were not excluded.

### Videographic Analysis

FL and VMV adapted a codebook from Haider et al.^6^ to capture encounter characteristics (initial or follow-up visit, hurriedness and duration of the encounter), to assess fidelity and contamination (whether and how the QBSAFE cards were used), and to assess its effect on the occurrence of domain-specific discussions and the degree of clinician responsiveness to the issues raised. FL and VMV trained a team of six additional reviewers (SB, SB, KG, CP, AvG, LV) to assess encounter recordings using the adapted codebook. No changes to these endpoints were made after the study started.

Fidelity required use of the QBSAFE cards in the first half of the clinical encounter. Reviewers coded each discussion observed as related to one or more of the QBSAFE issues (Quality of life, Burden of treatment, Safety, Avoidance of Future Events) or to metabolic management. These categories were further divided into subdomains. Quality of Life subdomains were based on the World Health Organization Quality of Life assessment instrument. Burden of treatment subdomains were based on the Treatment Burden Questionnaire.^7,8^ Safety discussions were subdivided into those about hypoglycemia and about other adverse effects of medications.

Avoidance of Future Events discussions were related to the prevention or management of T2D-related complications and were subdivided into discussions about either macrovascular or microvascular complications. Metabolic management discussions were subdivided into conversations regarding glycemic management, weight, lipids, and blood pressure management. Further description of each subdomain can be found in Table 1. When multiple discussions for a particular subdomain occurred, reviewers chose the one with which the clinician engaged to the greatest degree: from ignored, to only acknowledged, to responding with a revised care plan.

**Table 1.** Examples of Domain and Subdomains.

| Domain | Subdomain | Example |
| --- | --- | --- |
| Quality of Life | Physical | Pain, discomfort, energy, fatigue, sleep |
|  | Psychological | Positive/negative feelings thinking, learning, memory/<br>concentration, self-esteem |
|  | Level of Independence | Mobility, activities of daily living, dependence on<br>medication, work capacity |
|  | Social Relationships | Personal relationships, social support, sexual activity |
|  | Environment | Physical safety, home environment, financial resources,<br>opportunities for information, recreation, new skills,<br>pollution/noise/climate |
|  | Spirituality/Religion | Personal beliefs, structure to experience, coping with<br>difficulties through beliefs |
| Burden of Treatment | Administering or taking<br>medications | Injections, taking pills |
|  | Monitoring | Lab tests, imaging, blood pressure/sugar checks, time<br>spent or inconvenience of self-monitoring |
|  | Cost of medications,<br>monitoring, healthcare in<br>general | Concerns about cost, not being able to pay for services,<br>prescriptions |
|  | Scheduling/finding or<br>transportation to healthcare | Appointment times interfere with work, difficulty getting<br>to appointments, pharmacy |
|  | Administrative | Insurance, prior authorizations, pharmacy |
|  | Relationship with provider/others | Not a good patient/clinician relationship, not feeling listened to |
|  | Non-pharmacological interventions | Diet modifications, smoking/alcohol/drug cessation |
| Safety | Hypoglycemia | Real, fear of, prevention of |
|  | Other adverse events | Side effects directly attributable to diabetes medication |
| Avoidance of Future Events | Macrovascular | Heart, blood vessels |
|  | Microvascular | Kidneys, eyes, nerves |
| Metabolic Management | Glycemic Control | HbA1c, time in range, blood sugar levels |
|  | Lipids | Cholesterol/triglycerides |
|  | Blood Pressure | Monitoring/concerns |
|  | Weight | Gain/loss, concerns |

New care plans could include ordering additional testing, referrals, changing treatment, monitoring, or follow-up plans. Only one discussion per subdomain was evaluated. Reviewers captured how clinicians responded to discussions that reviewers judged as demanding clinical action. The full codebook can be found in Supplemental Material 2.

After an initial version of the codebook was completed, reviewers were trained and calibrated to ensure consistent measurement across the review team. Two review team members (FL and VM5) generated Gold Standard reviews by reviewing a set of encounter videos independently and meeting to resolve differences between them with assistance from an experienced encounter researcher and diabetologist (VMM). The other researchers were then asked to review videos, compare their results with the reference set, and discuss differences with the whole team, seeking both improvements to the codebook instructions and calibration between reviewers. The review team was considered calibrated when they achieved ≥70% raw agreement on domain-specific discussions.

After achieving calibration, reviewers reviewed encounter videos independently. Calibration was reassessed weekly by having all reviewers independently review an encounter video and comparing it against a reference review. If agreement fell below 70%, independent video review was stopped until reviewers could achieve ≥70% agreement for two consecutive videos.

Although reviewers were blinded to the trial allocation of their assigned encounters, they were able to discern if the QBSAFE cards were used in an encounter or not. Data was extracted into and managed using a REDcap database.^9,10^

### Statistical Analysis

Descriptive statistics were used to describe the study cohort and video review results with mean and standard deviations reported for continuous variables and counts and frequencies for categorical variables. Comparisons between arms were conducted by a linear mixed model with fixed effect of arm and cluster effect of clinician, reporting the p-value for the F-statistic. For binary outcomes, we used a generalized linear mixed model with a logit link and binary distribution with a fixed effect of arm and a random effect of clinician. For outcomes with more than 2 levels and ordinal in nature, the multinomial distribution and cumulative logit link was used, and if the categorical outcome is nominal then the generalized logit was used. All p-values reported are two-sided, with values <0·05 considered statistically significant. All statistical analyses were performed using SAS version 9·4 (SAS Institute Inc., Cary, NC).

## RESULTS

The trial enrolled and randomized 11 clinicians, 6 to the QBSAFE arm and 5 to the control arm. A total of 155 patients with appointments with these clinicians were enrolled in the trial; 12 (7·7%) were excluded post-enrollment due to an updated HbA1c ≤8% at the index visit, after consent. Six (3·8%) encounters had unusable videos, leaving 137 encounters with evaluable videos, 72 depicting usual care with the QBSAFE cards and 65 with usual care alone.

Table 2 reports baseline demographics and encounter characteristics. All baseline variables were similar between groups.

**Table 2.** Baseline characteristics of patients and encounters included in analysis.

|  | Arm |  |  |  |
| --- | --- | --- | --- | --- |
|  | Usual Care<br>(N=65) | QBSafe Cards<br>(N=72) | Total<br>(N=137) | P-value |
| **Sex, n (%)** |  |  |  | 0·6<sup>1</sup> |
| Female | 42 (65%) | 43 (60%) | 85 (62%) |  |
| Male | 23 (35%) | 29 (40%) | 52 (38%) |  |
| **Age, Mean (SD)** | 53·2 (10·8) | 58·5 (12·4) | 56·0 (11·9) | 0·14<sup>2</sup> |
| **Race, n (%)** |  |  |  | 0·07<sup>3</sup> |
| White | 16 (25%) | 31 (43%) | 47 (34%) |  |
| Black | 14 (22%) | 13 (18%) | 27 (20%) |  |
| Other | 29 (45%) | 24 (33%) | 53 (39%) |  |
| Unknown | 6 (9%) | 4 (6%) | 10 (7%) |  |
| **Ethnicity, n (%)** |  |  |  | 0·22<sup>1</sup> |
| Hispanic or Latino | 52 (80%) | 50 (69%) | 102 (74%) |  |
| Not Hispanic or Latino | 13 (20%) | 22 (31%) | 35 (26%) |  |
| **BMI, Mean (SD)** | 31·6 (5·1) | 30·9 (5·7) | 31·3 (5·4) | 0·61<sup>2</sup> |
| **HbA1c, Mean (SD)** | 10·2 (1·8) | 9·8 (1·7) | 10·0 (1·8) | 0·17<sup>2</sup> |
| **Primary Language, n (%)** |  |  |  | 0·064<sup>1</sup> |
| English | 18 (28%) | 32 (44%) | 50 (36%) |  |
| Spanish | 47 (72%) | 40 (56%) | 87 (64%) |  |
| **Was an interpreter present?, n (%)** |  |  |  | 0·46<sup>3</sup> |
| Yes, on the phone | 14 (22%) | 8 (11%) | 22 (16%) |  |
| Yes, in person | 2 (3%) | 5 (7%) | 7 (5%) |  |
| No | 49 (75%) | 59 (82%) | 108 (79%) |  |
| **Was a guest present?, n (%)** |  |  |  | 0·42<sup>1</sup> |
| No | 54 (83%) | 62 (86%) | 116 (85%) |  |
| Yes | 11 (17%) | 10 (14%) | 21 (15%) |  |
| **Length of video, Mean (SD)** | 22·2 (7·3) | 22·1 (6·9) | 22·1 (7·1) | 0·7<sup>2</sup> |
| **Patients usual clinician?, n (%)** |  |  |  | 0·43<sup>3</sup> |
| Clearly yes | 31 (48%) | 39 (54%) | 70 (51%) |  |
| Likely yes | 17 (26%) | 18 (25%) | 35 (26%) |  |
| Unclear | 9 (14%) | 8 (11%) | 17 (12%) |  |
| Likely no | 6 (9%) | 4 (6%) | 10 (7%) |  |
| Clearly no | 2 (3%) | 3 (4%) | 5 (4%) |  |
|  | Usual Care<br>(N=65) | QBSafe Cards<br>(N=72) | Total<br>(N=137) | P-value |
| **Did the encounter seem hurried?,**<br>**n (%)** |  |  |  | 0.05<sup>3</sup> |
| Mostly hurried | 6 (9%) | 1 (1%) | 7 (5%) |  |
| Mixed | 13 (20%) | 9 (13%) | 22 (16%) |  |
| Mostly unhurried | 46 (71%) | 62 (86%) | 108 (79%) |  |
| **How was the QBSafe tool**<br>**accessed?, n (%)** |  |  |  | ~ |
| Electronic | 0 (0%) | 1 (1%) | 1 (1%) |  |
| Paper/physical | 0 (0%) | 63 (88%) | 63 (46%) |  |
| Unclear | 0 (0%) | 6 (8%) | 6 (4%) |  |
| Not used | 65 (100%) | 2 (3%) | 67 (49%) |  |

<sup>1</sup>Generalized linear mixed model (GLMM) with a logit link and binary distribution for a comparison of the arm, reporting the p-value for the F-test; <sup>2</sup>Linear mixed-effects model to compare between arms account for clustering of clinician, reporting the p-value for the F-test; <sup>3</sup>Generalized linear mixed model (GLMM) with a multinomial distribution and cumulative logit link or generalized logit if the categorical outcome is nominal was used compare the arms, with F-statistic p-value being reported.

### Encounter Characteristics

Encounter length did not differ significantly between arms (22·2 vs. 22·1 minutes, p=0·698). Reviewers categorized 86% of QBSAFE arm encounters as unhurried compared with 71% of usual care encounters (*p*=0·045).

### Card usage and fidelity assessment

Reviewers did not observe any QBSAFE card use in the usual care arm. The mean number of cards selected in QBSAFE encounters was 2·2 (SD 1·2, Median 3 [IQR: 1-3]), with 8 of 72 (11·11%) patients selecting no cards and 41 (56·9%) selecting the maximum of 3. Clinicians almost always initiated discussion using the cards before the patients mentioned them (n=67, 93%). The cards were used as intended in 85% (n=61) of encounters (initiated by clinician in the first half of the encounter).

The five most frequently selected cards were: “I struggle with monitoring my blood sugar” (n=24), “I find it hard to follow your suggestions about diet and exercise” (n=20), “I have felt moments of pride while managing my diabetes” (n=20), “I struggle with remembering taking or managing my medications” (n=16), and “I would benefit from more help managing my diabetes” (n=15). The least chosen card was “I have something I would like to share with you but I know you probably will not be able to do much about it” (n=3) (Figure 1).

**Fig. 1:**
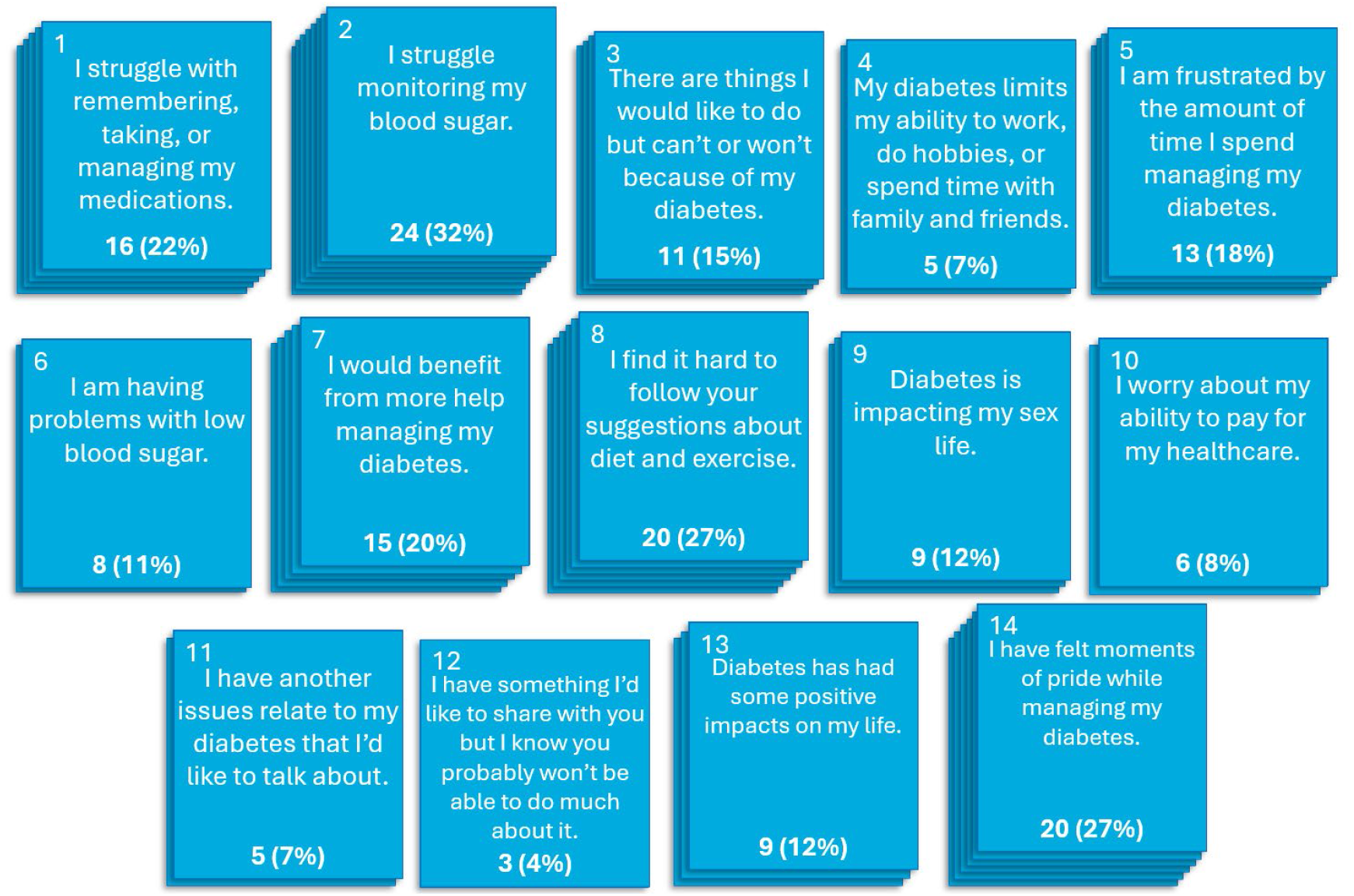
QBSAFE cards and their frequency of use in the intervention arm, number of encounters.

### Domain-specific discussions

The results of the intention-to-treat analysis are shown in Table 3. At the domain level, QBSAFE encounters had more discussions related to burden of treatment compared to the usual care arm. At the subdomain level, QBAFE encounters stimulated more discussions about the patient’s social risk (housing security, transportation) and about the burden associated with medications and with non-pharmacological interventions. The overall number of discussions were similar across arms for the remaining domains and subdomains, including metabolic management (*p*=0·40). Per protocol analysis showed identical results and is not included here.

**Table 3.** Occurrence of domain-specific discussions by study arm.

| Domain discussed in encounter | Arm |  |  |  |
| --- | --- | --- | --- | --- |
|  | Usual Care<br>(N=65) | QBSAFE Cards<br>(N=72) | Total<br>(N=137) | P-value |
| **Quality of Life, n (%)** | 51 (78.5%) | 62 (86.1%) | 113 (82.5%) | 0.37<sup>1</sup> |
| **Physical, n (%)** | 43 (66.2%) | 41 (56.9%) | 84 (61.3%) | 0.28<sup>1</sup> |
| **Psychological, n (%)** | 21 (32.3%) | 35 (48.6%) | 56 (40.9%) | 0.07<sup>1</sup> |
| **Level of independence, n (%)** | 6 (9.2%) | 12 (16.7%) | 18 (13.1%) | 0.24<sup>1</sup> |
| **Environment, n (%)** | 2 (3.1%) | 10 (13.9%) | 12 (8.8%) | **0.04<sup>1</sup>** |
| **Social relationships, n (%)** | 6 (9.2%) | 15 (20.8%) | 21 (15.3%) | 0.07<sup>1</sup> |
| **Religion, Spirituality, and Personal Beliefs, n (%)** | 1 (1.5%) | 4 (5.6%) | 5 (3.6%) | 0.18<sup>1</sup> |
| **Burden of Treatment, n (%)** | 41 (63.1%) | 59 (81.9%) | 100 (73.0%) | **0.02<sup>1</sup>** |
| **Related to medications, n (%)** | 11 (16.9%) | 33 (45.8%) | 44 (32.1%) | **0.0008<sup>1</sup>** |
| Relationship with provider/others, n (%) | 4 (6.2%) | 7 (9.7%) | 11 (8.0%) | 0.45<sup>1</sup> |
| Related to monitoring, n (%) | 16 (24.6%) | 18 (25.0%) | 34 (24.8%) | 0.95<sup>1</sup> |
| Costs of medications, monitoring, or<br>healthcare in general, n (%) | 7 (10.8%) | 5 (6.9%) | 12 (8.8%) | 0.43<sup>1</sup> |
| Clinician or lab visits, n (%) | 2 (3.1%) | 6 (8.3%) | 8 (5.8%) | 0.21<sup>1</sup> |
| Administrative burden, n (%) | 20 (30.8%) | 16 (22.2%) | 36 (26.3%) | 0.26<sup>1</sup> |
| Related to non-pharmacological<br>interventions or treatment, n (%) | 17 (26.2%) | 33 (45.8%) | 50 (36.5%) | **0.04<sup>1</sup>** |
| Safety, n (%) | 36 (55.4%) | 36 (50.0%) | 72 (52.6%) | 0.54<sup>1</sup> |
| Hypoglycemia, n (%) | 16 (24.6%) | 24 (33.3%) | 40 (29.2%) | 0.62<sup>1</sup> |
| Other adverse effects/harms, n (%) | 23 (35.4%) | 21 (29.2%) | 44 (32.1%) | 0.44<sup>1</sup> |
| Avoidance of future events, n (%) | 15 (23.1%) | 27 (37.5%) | 42 (30.7%) | 0.23<sup>1</sup> |
| Macrovascular, n (%) | 3 (4.6%) | 8 (11.1%) | 11 (8.0%) | 0.20<sup>1</sup> |
| Microvascular, n (%) | 14 (21.5%) | 22 (30.6%) | 36 (26.3%) | 0.43<sup>1</sup> |
| Metabolic management, n (%) | 59 (90.8%) | 62 (86.1%) | 121 (88.3%) | 0.40<sup>1</sup> |
| Glycemia, n (%) | 56 (86.2%) | 59 (81.9%) | 115 (83.9%) | 0.51<sup>1</sup> |
| Lipids, n (%) | 14 (21.5%) | 8 (11.1%) | 22 (16.1%) | 0.09<sup>1</sup> |
| Blood Pressure, n (%) | 10 (15.4%) | 15 (20.8%) | 25 (18.2%) | 0.41<sup>1</sup> |
| Weight, n (%) | 12 (18.5%) | 11 (15.3%) | 23 (16.8%) | 0.66<sup>1</sup> |
| **Encounters with new care plans, per domain**<br>**discussed** |  |  |  |  |
| Quality of life | 32 (49.2%) | 39 (54.1%) | 71 (51.8%) | 0.56<sup>1</sup> |
| Burden of treatment | 29 (44.6%) | 42 (58.3%) | 71 (51.8%) | 0.11<sup>1</sup> |
| Safety | 18 (27.7%) | 12 (16.7%) | 30 (21.8%) | 0.12<sup>1</sup> |
| Avoidance of future events | 9 (13.8%) | 17 (23.6%) | 26 (18.9%) | 0.20<sup>1</sup> |
| Metabolic management | 46 (70.7%) | 49 (68.1%) | 95 (69.3%) | 0.73<sup>1</sup> |
| <sup>1</sup>Generalized linear mixed model (GLMM) with a logit link and binary distribution for a comparison of the<br>arm adjusting for clustering effect of clinician, reporting the p-value for the F-test; |  |  |  |  |

### Responses to discussions

All encounters across both arms had at least one instance of a discussion that required action and at least one new care plan. Most encounters in the QBSAFE arm had a discussion about either quality of life (n=39, 54%) or burden of treatment that resulted in a new plan, 39 (54·1%) and (n=42, 58%). Most discussions about metabolic management in both the QBSAFE (n=46, 71%) and usual care (n=49, 68%) arms resulted in a new care plan. There was no difference in the prevalence of new plans between arms (Table 3)

## DISCUSSION

This study evaluated the effect of using novel QBSAFE cards on the content of primary care encounters with people living with T2D in a federally qualified health center. Clinicians were able to use the cards as intended, without lengthening the appointment compared to usual care. Compared to usual care, videographic analysis found that using QBSAFE cards contributed to more unhurried conversations about treatment burden and social risks without displacing discussions about metabolic management.

Our findings align with the results of other studies using conversation cards. In a pilot trial investigating the QBSAFE cards, 64% of patients reported that the QBSAFE cards facilitated conversations about their health.^11^ In a separate study that tested agenda-setting conversation cards in annual diabetes visits, most patients found the cards helpful for bringing up issues and concerns.^12^ These results suggest that integrating the QBSAFE cards into routine care may enhance communication and foster more meaningful, patient-focused interactions in T2D management without increasing the length of the encounter.^13^ These types of conversations are crucial for providing thoughtful and compassionate care that addresses the needs of both patients and clinicians.

Prior studies have also demonstrated the effectiveness of structured conversation tools such as question prompt lists (QPL) which enhance patient engagement and participation by encouraging patients to explore concerns they may otherwise withhold.^14-16^ Dimoska et al.^17^ found that QPLs empowered patients to initiate important discussions about quality of life, a finding mirrored in the current study. They also found mixed evidence on QPL effects on encounter duration.

However, a key difference is QPLs mostly support information gathering by enabling directed questions and answers, enabling an in-depth exploration of a particular topic, whereas conversation cards like QBSAFE support the identification and prioritization of different topics and stimulate more interactive dialogue between patients and clinicians.

Previous studies and clinical practice guidelines have also highlighted the importance of expanding clinical conversations beyond HbA1c to better address the holistic needs of diabetes patients.^6,18^ The current study suggests that the QBSAFE cards stimulate broader conversations on treatment burden, covering a wider spectrum of patient concerns without compromising or displacing conversations about metabolic management, enabling clinicians to respond to a wider range of patient needs. In particular, conversations about treatment burden are crucial in diabetes management, where the complexity of treatment plans and the emotional toll of managing a chronic condition can significantly affect quality of life and capacity to implement the care plans.^19^

Non-pharmacological interventions, such as diet changes and exercise, have been shown to improve glycemic management and overall health outcomes; they are also considered among the most burdensome aspects of care.^20^ By encouraging these discussions, the QBSAFE intervention can help surface a wider range of patients’ questions and concerns, allowing patients and clinicians to work together to address them. This approach is in line with recommendations that emphasize the importance of addressing non-medical factors in managing chronic conditions like type 2 diabetes.^21^

QBSAFE cards included statements intended to uncover problems, but also statements intended to reflect upon strengths and successes in a patient’s life with T2D. Encouraging patients to discuss successes and share pride in their diabetes management during clinical encounters is likely to result in more than just a “feel-good” moment—it may also reinforce behavioral changes, support self-efficacy and reveal untapped strengths, but more exploration is needed to determine this conclusively. It may also encourage patients to tell their stories, similarly to a narrative medicine approach, which may support communication, relationship-building, and more empathetic person-centered care.^22,23^ The card discussing moments of pride in diabetes management was the second most used card, consistent with the notion that patients are actively looking for opportunities to highlight their successes in their ongoing care.^24^

The QBSAFE cards resulted in more unhurried encounters without an increase in encounter duration. Unhurried conversations facilitate active listening and the establishment of therapeutic relationships, which are key to patient-centered communication, an essential aspect of managing chronic conditions, including diabetes.^25,26^

While we observed a difference in the types of conversations patients and clinicians had in the two study arms, this difference was small and only apparent in a few domains. Future work could aim to modify the QBSAFE cards to more effectively stimulate patient-centered discussions.

This modification could also aim to make some cards more responsive to patients’ needs; some cards were chosen only a handful of times, namely cards 4, 10, 11, and 12. Additionally, future work could examine how the use of the cards and their effect changes when used iteratively across multiple visits and with different members of the care team.

Additionally, the increase in the occurrence of discussions did not translate into the formation of new care plans. This suggests that, while interventions such as conversation cards may enable or initiate discussions on patient-centered domains, further resources that support the exploration of these topics and other elements beyond clinically focused decision-making may assist in leading to purposeful improvements to care plans. A potential avenue for further studies may be preparing and empowering clinicians to respond to issues raised by the cards, perhaps with information sheets^27^ or quick reference guides detailing available resources.

This study has some limitations. The study was conducted in a predominantly Spanish speaking, vulnerable population in need of interventions to achieve care that fits. While the availability of the cards in both English and Spanish allows for a broader population, this study was conducted in a single center, which limits the applicability of our findings to other primary care settings.

While it is possible that awareness of being videorecorded could have influenced behavior, a number of studies examining recorded encounters, video or otherwise, have demonstrated that this is not the case.^28-30^ Due to technical factors, some videos had poor quality audio, making some discussions difficult to understand, while several videos were not evaluable. Because it was not possible to blind reviewers to trial allocation–they could see the use of the cards during the encounter–this could have introduced bias. Only one discussion per domain was analyzed per encounter, selecting the discussion with more intensive clinician response, which may have overlooked other relevant discussions in the same domain in which, for example, clinicians may have addressed concerns without the need for a new plan. This was done to bias our assessments to give the maximum credit to clinicians. Variability in how reviewers assessed areas with fewer discussions, such as quality of life related to spirituality or social relationships, reduces their reliability. While both English and Spanish-speaking patients were included, sample size was insufficient to detect any differences in how those groups used QBSAFE cards. The impact of the QBSAFE cards on encounters with interpreters also remains unclear. Future work should expand the sample to enable the assessment of possible interactions between the effectiveness of the intervention and these language factors. Further study of clinician workload and satisfaction could also help assess whether such tools are sustainable in practice.

The QBSAFE cards facilitated more frequent and nuanced discussions about quality of life, treatment burden, and non-pharmacological strategies often underrepresented in routine practice.

They created space for patients to voice priorities that extended beyond glycemic control, supporting more individualized, person-centered conversations. These findings suggest that low-burden, structured communication tools can shift clinical dialogue toward shared decision-making and contextualized care, even within time-limited settings. Future research should explore the scalability of QBSAFE across diverse populations and examine its long-term impact on patient outcomes, clinician experience, and health system integration.

## Data Availability

Individual participant data that underlie the results reported in this Article will be available from the corresponding author on reasonable request. Participant video recordings cannot be shared due to confidentiality issues.

## Contributors

All authors had full access to all the data in the study and had final responsibility for the decision to submit for publication.

## Acknowledgement

This study was funded by NIDDK R01DK129616.

## References

1. Nowakowska M, Zghebi SS, Ashcroft DM, et al. The comorbidity burden of type 2 diabetes mellitus: patterns, clusters and predictions from a large English primary care cohort. BMC Medicine 2019; 17(1): 145.

2. Kunneman M, Griffioen IPM, Labrie NHM, Kristiansen M, Montori VM, van Beusekom MM. Making care fit manifesto. BMJ Evidence-Based Medicine 2023; 28(1): 5–6.

3. Stacey D, Lewis KB, Smith M, et al. Decision aids for people facing health treatment or screening decisions. Cochrane Database Syst Rev 2024; 1(1): Cd001431.

4. Goge S, Tran C, Lewis KB, Carley M, Bennett C, Stacey D. What Is the Effectiveness of Type 2 Diabetes–related Patient Decision Aids? Secondary Analysis of a Systematic Review. Canadian Journal of Diabetes 2025.

5. Clark JE, Boehmer KR, Breslin M, et al. Quality of life, burden of treatment, safety, and avoidance of future events (QBSAfe) protocol: a pilot study testing an intervention to shift the paradigm of diabetes care. Pilot Feasibility Stud 2021; 7(1): 196.

6. Haider S, El Kawkgi O, Clark J, et al. Beyond hemoglobin A1c: a videographic analysis of conversations about quality of life and treatment burden during clinical encounters for diabetes care. Endocrine 2021; 73(3): 573–9.

7. Tran VT, Harrington M, Montori VM, Barnes C, Wicks P, Ravaud P. Adaptation and validation of the Treatment Burden Questionnaire (TBQ) in English using an internet platform. BMC Med 2014; 12: 109.

8. Organization WH. Programme on Mental Health: WHOQOL. 2012.

9. Harris PA, Taylor R, Minor BL, et al. The REDCap consortium: Building an international community of software platform partners. Journal of Biomedical Informatics 2019; 95: 103208.

10. Harris PA, Taylor R, Thielke R, Payne J, Gonzalez N, Conde JG. Research electronic data capture (REDCap)--a metadata-driven methodology and workflow process for providing translational research informatics support. J Biomed Inform 2009; 42(2): 377–81.

11. Haider S, Gonzalez-Lopez C, Clark J, et al. Feasibility and Acceptability of an Agenda-Setting Kit in the Care of People With Type 2 Diabetes: The QBSAFE ASK Feasibility Study. Clin Diabetes 2024; 42(3): 358–63.

12. Lomborg K, Munch L, Krøner FH, Elwyn G. “Less is more”: A design thinking approach to the development of the agenda-setting conversation cards for people with type 2 diabetes. PEC Innovation 2022; 1: 100097.

13. Elmore N, Burt J, Abel G, et al. Investigating the relationship between consultation length and patient experience: a cross-sectional study in primary care. Br J Gen Pract 2016; 66(653): e896–e903.

14. Eggly S, Hamel LM, Foster TS, et al. Randomized trial of a question prompt list to increase patient active participation during interactions with black patients and their oncologists. Patient Education and Counseling 2017; 100(5): 818–26.

15. Amundsen A, Bergvik S, Butow P, Tattersall MHN, Sørlie T, Nordøy T. Supporting doctor-patient communication: Providing a question prompt list and audio recording of the consultation as communication aids to outpatients in a cancer clinic. Patient Education and Counseling 2018; 101(9): 1594–600.

16. Coyne I, Sleath B, Surdey J, et al. Intervention to promote adolescents’ communication and engagement in diabetes clinic encounters: A pilot randomized controlled trial. Patient Educ Couns 2024; 126: 108322.

17. Dimoska A, Tattersall MH, Butow PN, Shepherd H, Kinnersley P. Can a “prompt list” empower cancer patients to ask relevant questions? Cancer 2008; 113(2): 225–37.

18. Committee ADAPP. 4. Comprehensive Medical Evaluation and Assessment of Comorbidities: Standards of Care in Diabetes—2025. Diabetes Care 2024; 48(Supplement_1): S59–S85.

19. Sagastume D, Siero I, Mertens E, Cottam J, Colizzi C, Peñalvo JL. The effectiveness of lifestyle interventions on type 2 diabetes and gestational diabetes incidence and cardiometabolic outcomes: A systematic review and meta-analysis of evidence from low- and middle-income countries. EClinicalMedicine 2022; 53: 101650.

20. Espinoza P, Varela CA, Vargas IE, et al. The burden of treatment in people living with type 2 diabetes: A qualitative study of patients and their primary care clinicians. PLoS One 2020; 15(10): e0241485.

21. Wang X, Kang J, Liu Q, Tong T, Quan H. Fighting Diabetes Mellitus: Pharmacological and Non-pharmacological Approaches. Curr Pharm Des 2020; 26(39): 4992–5001.

22. Drossman DA, Chang L, Deutsch JK, et al. A Review of the Evidence and Recommendations on Communication Skills and the Patient-Provider Relationship: A Rome Foundation Working Team Report. Gastroenterology 2021; 161(5): 1670-88.e7.

23. Charon R. The patient-physician relationship. Narrative medicine: a model for empathy, reflection, profession, and trust. Jama 2001; 286(15): 1897–902.

24. Robertson SM, Stanley MA, Cully JA, Naik AD. Positive emotional health and diabetes care: concepts, measurement, and clinical implications. Psychosomatics 2012; 53(1): 1–12.

25. Ballard DI, Mandhana DM, Tesfai Y, et al. Unhurried Conversations in Health Care Are More Important Than Ever: Identifying Key Communication Practices for Careful and Kind Care. Ann Fam Med 2024; 22(6): 533–8.

26. Berwick DM. What ‘patient-centered’ should mean: confessions of an extremist. Health Aff (Millwood) 2009; 28(4): w555–65.

27. Andermann A. Taking action on the social determinants of health in clinical practice: a framework for health professionals. Cmaj 2016; 188(17-18): E474–e83.

28. Pringle M, Stewart-Evans C. Does awareness of being video recorded affect doctors’ consultation behaviour? Br J Gen Pract 1990; 40(340): 455–8.

29. Penner L, Orom H, Albrecht T, Franks M, Moore T, Ruckdeschel J. Camera-Related Behaviors during Video Recorded Medical Interactions. Journal of Nonverbal Behavior 2007; 31: 99–117.

30. Henry SG, Jerant A, Iosif AM, Feldman MD, Cipri C, Kravitz RL. Analysis of threats to research validity introduced by audio recording clinic visits: Selection bias, Hawthorne effect, both, or neither? Patient Educ Couns 2015; 98(7): 849–56.

