## Supplemental Material 1 and 2 for "Videographic Analysis of an Intervention to Improve Patient-Centered Care for People Living with Type 2 Diabetes: the QBSAFE Randomized Trial"

### Supplementary Material 1. - CONSORT 2010 checklist of information to include when reporting a randomised trial*

| Section/Topic | Item No | Checklist item | Reported on page No |
|---|---|---|---|
| **Title and abstract** | | | |
| | 1a | Identification as a randomised trial in the title | Title Page |
| | 1b | Structured summary of trial design, methods, results, and conclusions (for specific guidance see CONSORT for abstracts) | 2 |
| **Introduction** | | | |
| Background and objectives | 2a | Scientific background and explanation of rationale | 4 |
| | 2b | Specific objectives or hypotheses | 5 |
| **Methods** | | | |
| Trial design | 3a | Description of trial design (such as parallel, factorial) including allocation ratio | 5 |
| | 3b | Important changes to methods after trial commencement (such as eligibility criteria), with reasons | 5 |
| Participants | 4a | Eligibility criteria for participants | 6 |
| | 4b | Settings and locations where the data were collected | 5 |
| Interventions | 5 | The interventions for each group with sufficient details to allow replication, including how and when they were actually administered | 6 |
| Outcomes | 6a | Completely defined pre-specified primary and secondary outcome measures, including how and when they were assessed | 7-8 |
| | 6b | Any changes to trial outcomes after the trial commenced, with reasons | 7-8 |
| Sample size | 7a | How sample size was determined | Defined in clinical outcomes manuscript. |
| | 7b | When applicable, explanation of any interim analyses and stopping guidelines | Not Applicable |
| Randomisation: | | | |
| Sequence generation | 8a | Method used to generate the random allocation sequence | 6 |
| | 8b | Type of randomisation; details of any restriction (such as blocking and block size) | 6 |
| Allocation concealment mechanism | 9 | Mechanism used to implement the random allocation sequence (such as sequentially numbered containers), describing any steps taken to conceal the sequence until interventions were assigned | 6 |

| | | | |
|---|---|---|---|
| Implementation | 10 | Who generated the random allocation sequence, who enrolled participants, and who assigned participants to interventions | 6 |
| Blinding | 11a | If done, who was blinded after assignment to interventions (for example, participants, care providers, those assessing outcomes) and how | 7 |
| | 11b | If relevant, description of the similarity of interventions | Not Applicable |
| Statistical methods | 12a | Statistical methods used to compare groups for primary and secondary outcomes | 9 |
| | 12b | Methods for additional analyses, such as subgroup analyses and adjusted analyses | 9 |
| **Results** | | | |
| Participant flow (a diagram is strongly recommended) | 13a | For each group, the numbers of participants who were randomly assigned, received intended treatment, and were analysed for the primary outcome | 10 |
| | 13b | For each group, losses and exclusions after randomisation, together with reasons | Defined in clinical outcomes manuscript. |
| Recruitment | 14a | Dates defining the periods of recruitment and follow-up | 6 |
| | 14b | Why the trial ended or was stopped | Not Applicable |
| Baseline data | 15 | A table showing baseline demographic and clinical characteristics for each group | 21 |
| Numbers analysed | 16 | For each group, number of participants (denominator) included in each analysis and whether the analysis was by original assigned groups | 10 |
| Outcomes and estimation | 17a | For each primary and secondary outcome, results for each group, and the estimated effect size and its precision (such as 95% confidence interval) | 10-11 |
| | 17b | For binary outcomes, presentation of both absolute and relative effect sizes is recommended | 10 |
| Ancillary analyses | 18 | Results of any other analyses performed, including subgroup analyses and adjusted analyses, distinguishing pre-specified from exploratory | 10-11 |
| Harms | 19 | All important harms or unintended effects in each group (for specific guidance see CONSORT for harms) | 11 |
| **Discussion** | | | |
| Limitations | 20 | Trial limitations, addressing sources of potential bias, imprecision, and, if relevant, multiplicity of analyses | 15 |
| Generalisability | 21 | Generalisability (external validity, applicability) of the trial findings | 15 |
| Interpretation | 22 | Interpretation consistent with results, balancing benefits and harms, and considering other relevant evidence | 13 |
| **Other information** | | | |

| | | | |
|---|---|---|---|
| Registration | 23 | Registration number and name of trial registry | 5 |
| Protocol | 24 | Where the full trial protocol can be accessed, if available | 5 |
| Funding | 25 | Sources of funding and other support (such as supply of drugs), role of funders | 3 |

Citation: Schulz KF, Altman DG, Moher D, for the CONSORT Group. CONSORT 2010 Statement: updated guidelines for reporting parallel group randomised trials. BMC Medicine. 2010;8:18.
© 2010 Schulz et al. This is an Open Access article distributed under the terms of the Creative Commons Attribution License (http://creativecommons.org/licenses/by/2.0), which permits unrestricted use, distribution, and reproduction in any medium, provided the original work is properly cited.

*We strongly recommend reading this statement in conjunction with the CONSORT 2010 Explanation and Elaboration for important clarifications on all the items. If relevant, we also recommend reading CONSORT extensions for cluster randomised trials, non-inferiority and equivalence trials, non-pharmacological treatments, herbal interventions, and pragmatic trials. Additional extensions are forthcoming: for those and for up-to-date references relevant to this checklist, see www.consort-statement.org.

**Supplemental Material 2. - Videographic Analysis Codebook**

**<u>Basic Information:</u>**

1. **Patient ID**: as noted within REDCAP

2. **Coder Name:**

3. **Type of recording:** Encounters will be coded as either be video or audio recording. A video encounter will be coded if either the clinician, patient, or both are visibly seen on the video recording. If the screen is black/blank with only audio present – code as an audio recording.
    a. Video
    b. Audio
    c. Mixed (e.g., started off as video and was converted to audio only)

4. **Length of encounter:** Length of encounter is from the time the clinician and patient are both present within the encounter. Do not include time while patient is waiting for the clinician to be present within the exam room. Time should be written as minutes: seconds. If Encounter A is made up of two videos (video 1 = 8:51 and video 2 = 8:51) then write total time of encounter (total length = 17 min and 42 sec; or 17:42). If the clinician leaves and returns, only count the time the clinician is in the room.

5. **Complete record:**
    a. Likely yes (recording includes the whole encounter)
    b. Unclear
    c. Likely no (recording stops before encounter ends)

6. **Usual clinician**: Was this the patient's usual clinician? Yes, if it is clear that the patient and clinician met before and refer to prior visits or conversations. No if this is the first visit.
    a. Clearly Yes
    b. Likely Yes
    c. Unclear
    d. Likely No
    e. Clearly No

7. **Hurriedness**: Did the encounter seem hurried? Mostly hurried: patient and clinician seemed rushed either physically (one of them never sits down or stops moving or multitasking), verbally (language pressure, without pause, never let the other speak, interrupts often to get to the point or conclusion), or nonverbally (signals impatience, to move on, signals to leave early), emotional expressions and small talk is severely curtailed, the encounter is interrupted. Mostly unhurried: patient and clinician keep an efficient but not rushed pace, there is turn taking and pauses in language, there is rest in their movement and bodies that lean forward to invite and maintain interaction, there are follow-up statements ("go on", "tell me more" "what else?") and curious questions ("why is that?" "what else is in your mind"?) that seek to extend or deepen the interaction, time is made for small talk, and emotion is expressed freely. There are minimal or inconsequential interruptions.
    a. Mostly hurried
    b. Mixed
    c. Mostly unhurried

**<u>QBSafe</u>**:

8. **Reference to QBSAFE tool**: Was the QBSAFE ASK tool referred to within the clinical encounter at least once? Code "yes" if either clinician or patient referred to the tool in any capacity during the encounter. This can be either the clinician/patient pointing to the tool, referencing the tool within their conversation, commenting about the tool, etc. Code "no" if the tool was ignored i.e. tool placed

aside and not referenced to physically or verbally during the clinical encounter when both clinician and patient are present.

a. Yes
b. Unclear
c. No

9. Did patient choose any QBSAFE ASK cards to be discussed during the clinical encounter? **Patients chose a QBSAFE card**: Only code what is present in the video/audio encounter. Code "yes" if patient said any of the cards applied to them during current encounter. This includes whether the patient chose a card and *adjusted* words present on card (example. Chose card and replaced *diabetes* with another condition/general medical problems). Code "no" if the patient said the cards did not apply to them – even if they reference an issue (without the use of the tool) that is addressed by the cards.

   a. Yes
   b. Unclear
   c. No

If YES to question 10, answer the following questions:

10. **Accessing the QBSAFE tool**: Tool will either be accessed electronically or physically. Electronically includes using the online website, providing an electronic copy of the tool (email, pdf, screenshot/screen-sharing, etc). If the tool was provided in the form of cards or paper handout, then code as "paper/physical". How was the QBSAFE tool accessed?

    a. Electronic
    b. Paper/physical
    c. Unclear

11. **Who brought up cards**: Who first brought up the QBSAFE ASK tool within the encounter?

a. Clinician

b. Patient

12. **Timing of cards**: When was the QBSAFE ASK tool introduced within the encounter?

    a. First half of the encounter (broadly, i.e., before the physical exam)

    b. Second half of the encounter (broadly, i.e., after the physical exam)

13. **Anything to note** about the introduction of the QBSAFE tool (in particular, any verbal comment by patient and/or clinician about the cards, their pertinence, value, or usefulness or lack thereof): examples may include "these cards really feel like they get me" or "I had to pick a card but I don't find these helpful."

14. **Describe non-verbal responses** of the clinician to the QBSAFE tool. If a non-verbal response is impossible to detect, say because the encounter is audio-only or if the clinician is not visible in the video, enter "could not be observed". Examples may include: smiled at cards, sighed and picked up cards, threw the cards, or received cards from patient and read them.

**Discussions**:

Complete the following for any discussion of a QBSafe domain.

A discussion contains the following:

1. There must be a verbal exchange between patient and clinician with minimum of a 1 utterance per participant.

    a. For example, the exchange (C: "Are you having problems sleeping?", P: "No") would satisfy this second rule.

    b. The exception to this rule is **if the patient is the only one contributing an utterance**. This is to capture instances where a patient brings something up but is ignored by the clinician.

2. There must be an expression of some expression of importance, investment, or that something is undesirable or desirable, and that the subject of that expression has either improved, ceased, worsened, or persisted. Example:

| Clinician Speech | Patient Speech | Result |
|---|---|---|
| "Are you having trouble sleeping?" | "No." | No problem or expression of interest |
| | "Yes"/"It's gotten worse in the past week" | Worse/new/persisting issue |
| | "Somewhat, but less than before."/"It's much better than before." | Improvement of an issue |

3. The patient and/or the clinician talk about something in a QBSafe domain.

Discussions do not have to be time contiguous. Ex: If a discussion is interwoven throughout the encounter, code this is as one discussion, not two unique discussions.

Only code once per subdomain, if discussions pertaining to a subdomain happen during the encounter. If multiple conversations/discussions occur under one subdomain, **please select a conversation in which the physician takes the most proactive steps to address a specific issue or problem.**

**Disclaimer:** The chosen conversation does not need to be the most clinically relevant to the encounter. Your selection should emphasize the physician's response, rather than the overall clinical importance of the issue discussed.

When coding discussions, it is important to pay attention to what the main point of the discussion is. A discussion may cross touch on several domains, but reviewers should attempt to identify the main point of the conversation and code that discussion by a specific domain.

1. **Was there a Discussion of at least one QBSafe domain?** If yes to at least one of them, choose among the subdomains the topics of discussion that were present (mark box):

    - Quality of Life (QoL)
        - Physical (pain & discomfort; energy & fatigue; sleep & rest)
        - Psychological (positive feelings; thinking, learning, memory & concentration; self-esteem; bodily image & appearance; negative feelings)
        - Level of Independence (mobility; activities of daily living; dependence on medication or treatment; work capacity).
        - Social Relationships (personal relationships; social support; sexual activity)
        - Environment (physical safety & security, home environment, financial resources, health & social care: accessibility and quality; Opportunities for acquiring new information and skills; Participation in and opportunities for recreation/ leisure activities; Physical environment (pollution/noise/traffic/climate; Transport)
        - Spirituality/Religion/ Personal Beliefs

    - Burden of Treatment
        - Related to taking or administering medications (insulin injections, taking pills)
        - Related to monitoring (frequency/time spent or inconvenience incurred self-monitoring, lab tests, imaging, blood sugar checks, weight, blood pressure, etc.)
        - Costs of medications, monitoring, or healthcare in general
        - Related to scheduling/finding and transporting/getting to lab visits or doctor's appointments.

        - Administrative burden (dealing with insurance, prior authorizations, pharmacy, or other)
        - Relationship with provider/others (not a good patient/physician relationship, not feeling listened to, feeling like a burden to others)
        - Related to non-pharmacological interventions or treatment (diet modifications, smoking/alcohol cessation, etc.)

    - Safety
        - Hypoglycemia (Real, perceived, fear of, prevention of)
        - Other adverse effects/harms of diabetes medication (Some mentions of adverse effects may fall into other domains (e.g., stomach pain could be coded as QOL-Physical). If the adverse effect can be directly attributed to diabetes medication, code here)

    - Avoidance of Future Events (monitoring/inspection of signs or symptoms, or testing)
        - Macrovascular
            - E.g., heart, vessels
        - Microvascular
            - E.g., kidneys, eyes, nerves

    - Metabolic Management
        - Glycemia (HbA1c, time in range, blood sugar levels)
        - Lipids
        - Blood pressure (monitoring or concerns/successes)
        - Weight

2. Did this discussion require action on the part of the clinician?

    a) Clearly Yes

b) Likely Yes

c) Unclear

d) Likely No

e) Clearly No

3. **IF YES TO THE ABOVE QUESTION**: How did the clinician respond to the issue/point of success? (response descriptions in appendix)

   a. Ignored/did not address

   b. Acknowledged without exploring or proposing a solution

   c. Acknowledged, explored causes and/or consequences, asked elaborating questions but did not explore changes.

   d. Acknowledged and explored causes and/or consequences, asked elaborating questions, responded by exploring changes but no changes were made.

   e. New plan in response:

      i. Proposed to monitor and/or observe

      ii. Ordered additional testing

      iii. Referred to specialist or other assistance (social work)

      iv. Changed medication regimen or treatment (new meds, diet, exercise)

      v. Changed frequency or method of monitoring

      vi. Changed frequency or method of follow up

      vii. Other (enter text)

**Clinician Response Types:** Assess how the clinician responded to the QBSafe issues that came up. Ignored and acknowledge are exclusive – meaning no other response to the issue could have been present. i.e. can not check “acknowledged” and “asked more questions” à pick “asked more questions” alone.

*Ignored:* Clinician did not address the issue(s) during the clinical encounter. If clinician referenced the issue later in the encounter code as “addressed”, or code clinician response as below.

*Acknowledged:* Clinician addressed or acknowledged the issue brought up. Clinician must refer to the issue but not offer any other response (as below). If clinicians reference the issue but recommend addressing at future visit – code as “monitor” instead of addressed.

*Asked more questions:* Clinician asked elaborating questions. This is not an exclusive response. If clinician asked more questions and suggested a behavioral change – code both.

*Monitor:* Clinician offers continuing to monitor symptoms, condition, etc and address at future visit/follow up.

*Ordered additional testing:* Clinician asks patient to complete blood work, diagnostic imaging, or other tests (e.g., cardiac stress test, sleep study).

*Referral:* Clinician offers referral to consultant or general practitioner to address patient’s concern. Code as “referral” if referral was offered and patient denied or already seeing said consultant.

*New or change in medication or treatment plan:* Clinician recommends changing, adjusting or offering new medication. Code if clinician recommends change in timing of medication, switching from a twice a day to once a day formulation, adjustments to dose of insulin/insulin pump.

*Behavioral change:* Clinician recommends behavioral change such as dietary or exercise changes. Code if clinician recommends the use of non-pharmacologic means to address patient’s concern (i.e. use of pill box, placing medication on dining table).

*Change in monitoring (of diabetes):* Clinician recommend change in glucose monitoring. Code if clinician recommends switching to CGM, change in the number of glucose checks, etc.

*Change in follow up with clinician:* Clinician recommends change in follow up with themselves, ancillary staff, or seeing an established provider sooner/later.
